# Preferential Publication Bias in Nepal’s Medical Journals

**DOI:** 10.1101/2022.08.15.22278758

**Authors:** Dipendra Prasad Pant, Bikram Acharya, Mukunda Raj Kattel

**Affiliations:** Policy Research Institute, Kathmandu Nepal

**Keywords:** Preferential publishing, self-publishing, Medical Journals, Nepal, publication bias

## Abstract

The article has explored the preferential publication bias of Nepal’s medical journals. To this end, it has reviewed the frequency and proportion of preferential publishing and editorial involvement in endogenous publication practices in six national medical journals registered in the Scopus database system. For the analysis of the data, social network analysis application – VOSviewer for graphical visualisation has been used. Editorial engagement in self-publishing and preferential publishing is found to be common in all journals. The study suggests that as long as the trend of preferential and sequestered publication continues, the integrity and validity generated and disseminated by the journals risks losing trust by the community concerned and the chances of these non-mainstream journals contributing to mainstream journals being slim. And, by way of recommendatory conclusions, it offers the following four questions and areas for further investigation to arrive at the clarity and understanding of some of the issues that have been flagged in the findings and discussions: (a) Why do editors engage in excess self-promotion using the outlet they are supposed to objectively and transparently manage? (b) What is the motivating factor for authors to rely on a particular journal to get published despite having multiple outlets to pick and choose from? (c) Why should the editors and reviewers of scientific contributions maintain the networking silo to include a few in the loop and exclude others from it?

## Introduction

Ideally, scientific journal papers should be free from any kinds of biases. In practice, however, publication biases get reflected in various forms and degrees. Some studies get never published due to “file-drawer problem” – a situation whereby completed studies are never published. Generally, studies that are statistically significant (Nair, 2019), that explore new and immediate social issues into the academic discussion, and those with positive results get published with priorities (Mlinarić et al., 2017). Trials with positive results are published sooner than other trials (Hopewell et al., 2001). Time-lag bias, specifically in the case of health and clinical studies, results in due to various reasons such as the nature of results of the studies and the process duration of regulatory and scrutiny authorities.

Individual papers are generally believed to be free from publication biases as they are polished through editorial vetting, reviews and scrutiny. The process makes the papers more scientific, less biased and trustworthy than those that avoid or escape such scrutiny (Childe, 2006). For such qualities, editors serve both as quality controllers and gatekeepers. They, in principle, work towards avoiding all forms of biases via the application of established editorial norms, such as fairness, objectivity, transparency, and rigour. However, some journals may not follow rigorous processes of publication practice, ultimately allowing a situation whereby publication biases creep into due to omissions, lapses and unethical practices on the part of editors and publishers. The journals that publish manuscripts without a rigorous process of scrutiny risk endorsing what Anderson and Rainie (2017) call a doctored narrative, which poses a serious threat to scientific inquiries and their outcomes. In such a case, the journals cannot play a significant role in advancing scientific knowledge rather they become the platform for unverified or doubtful scientific knowledge.

Editor-authorship is one of the forms of publication bias as editor-authors exert undue influences on their colleagues to sympathetically review their submissions and clear them for publication (Luty et al., 2009). Such contributions are, however, highly selective, purposefully invited and strategically aimed. Contributing multiple full-length articles regularly by the editors having an executive role in the publication processes of journal articles contradicts the established editorial norms(Luty et al., 2009; Mani et al., 2013), which should not be breached to keep the integrity and standing of the journals. Such a tendency compromises the integrity of journals, breaches the trust of contributors, promotes self-adulation and, consequently, does a huge disservice to scientific enquiry (Bošnjak et al., 2011; Teixeira da Silva, 2017).

As a network of authors, co-authorship is one of the measures to analyse an individual scholar’s ability to collaborate with others and explore knowledge from different perspectives within a similar cross-discipline domain of scientific enquiry. Authors’ field of academic study, the level of pressure to get published, ranks in academia and the pursuit of quality are probable factors to bring authors together for co-authorships (Piette & Ross, 1992). Scholars contributing to specific research areas and those working in different geographical locations tend to participate in specific co-authorship networks (Uddin et al., 2012), which provide a platform for different sets of talent to jointly or collectively produce a research output (Kumar, 2015). Co-authorship, however, is not always welcome as it may do diservice to ethical publication also constituting sometimes to publication bias. The net relationship between the outputs attributable to individual author after discounting for the number of authors and the co-authorship is negative (Hollis, 2001). Also, unjustified and prolific co-authorship simply works as one of the ways for inflation of scientometric indices that do not necessarily further reflect the true quality of an individual’s research output (Masic & Jankovic, 2021).

Editors and publishers may also create publication bias by preferring the work of only one or a few authors, allowing to publish in the journals they edit and similar other ways. If every journal in specific national contexts allows biased co-authorship, editor-authorship or unusual preference to a single author such practices could qualify a publication bias thereby posing a serious problem to the national scientific inquiries and studies. The practice ultimately becomes one of the reasons for national publications of not excelling internationally and maintaining international standards.

In this background, this article explores the status of Nepal’s journals vis-à-vis maintaining and achieving international quality standards. For the purpose, we explore the extent of editorial and publishers’ practices specifically the preferential publication biases of Nepal’s medical journals, which exist to serve Nepal’s medical field of research and inquiries confined in small communities of practice. This article, therefore, is believed to contribute to stirring contributors and publishers of Nepali journals to reach out to international communities of practice for quality research and publication besides contributing to bridging national researchers with the internal practice and communities.

## Methodology

For this paper, we analysed full-length articles published in the Nepali medical journals indexed in the Scopus database: the Journal of Nepal Medical Association (JNMA), the Journal of Nepal Paediatric Society (JNPS), Kathmandu University Medical Journal (KUMJ), the Journal of Nepal Health Research Council (JNHRC), Nepal Medical College Journal (NMCJ) and Nepal Journal of Ophthalmology (NJO). The journals are among the 238 journals indexed by Nepal Journal Online (NepJol), the only academic journal database in Nepal managed by Tribhuvan University Central Library. The six journals are the only Nepalese journals ever indexed in the Scopus database (two of them have now been removed from the indexing). However, we used the data of the removed journal of until the date they were active under the Scopus database.

**Table 1:** Nepal’s Medical Journals Indexed by Scopus Database

| Name of Journal | Quartile performance | Discipline | Publisher | H-Index of journals | Number of articles indexed in Scopus Database | Remarks |
| --- | --- | --- | --- | --- | --- | --- |
| Journal of the Nepal Medical Association (https://www.scimagojr.com/journalsearch.php?q=15831&tip=sid&clean=0) | 1999 (Q3), 2000-2005 (non included), 2006 (Q4), 2007-2015 (Q3), 2016-2020 (Q4) | Medicine (Miscellaneous) | Nepal Medical Association | 19 | 1338 | Currently indexed in Scopus |
| Journal of Nepal Health Research Council (https://www.scimagojr.com/journalsearch.php?q=21100197340&tip=sid&clean=0) | 2011 (Q4), 2012-2016(Q3), 2017 (Q4), 2018-2020 (Q3) | Medicine (Miscellaneous) | Nepal Health Research Council | 13 | 720 | Currently Indexed in Scopus |
| Kathmandu University Medical Journal<br>(https://www.scimagojr.com/journalsearch.php?q=4000148103&tip=sid&clean=0) | 2004(Q3), 2005(Q4)<br>2005-2019(Q3), 2020 (Q4) | Medicine (miscellaneous) | Kathmandu University (In association with medical colleges affiliated with Kathmandu University) | 25 | 1361 | Currently indexed in Scopus |
| Journal of Nepal Paediatric Society<br>(https://www.scimagojr.com/journalsearch.php?q=19700174963&tip=sid&clean=0) | 2009-2010(Q4),<br>2011-2014 (Q3),<br>2015-2020(Q4) | Pediatrics, Perinatology and Child Health | Nepal Paediatric Society | 8 | 257 | Currently indexed in Scopus |
| Nepal Medical College journal: NMCJ<br>(https://www.scimagojr.com/journalsearch.php?q=145147&tip=sid&clean=0) | 2004-2005(Q4),<br>2006-2017 (Q3) | Medicine (miscellaneous) | Nepal Medical College | 21 | 444 | Discontinued in Scopus |
| Journal Of The Nepal Ophthalmic Society<br>(https://www.scimagojr.com/journalsearch.php?q=19900191831&tip=sid&clean=0) | 2010-2011 (Q4)<br>2012-2016(Q3)<br>2017 (Q4)<br>2018 (Q3)<br>2019 (Q4) | Medicine<br>Medicine (miscellaneous) | Nepal Ophthalmic Society | 13 | 92 | Discontinued |

The Scopus database is one the largest bibliometric database maintained by Elsevier and has more than 22,000 active titles besides some 13,000 inactive ones. We used Scopus database as the source because of its global recognition as one of the quality indexing systems. The Scopus database publicly provides journal articles related information and allows downloading the articles with the help of a specified script via an inbuilt search engine. Indexed under it, journals get more visible, enjoy more possibilities of availing themselves in the library databases and, reach out to a broader academic audience. It, however, does not mean that getting indexed under Scopus is the ultimate achievement, as there are other indexing databases.

To search the data for the study, we used the search code “AFFILCOUNTRY(Nepal)” in the Scopus search engine, collected the name of the Nepali journals indexed and used the individual search code for each journal with EXACTSRCTITLE (“*Name of Nepalese Journal*”) AND (LIMIT-TO (DOCTYPE, “ar”). The complete search code for particular journal is (AFFILCOUNTRY (nepal) AND ((PUBYEAR > 1980) AND (PUBYEAR < 2022))) AND (LIMIT-TO (SRCTYPE, “j”)) AND (LIMIT-TO (DOCTYPE, “ar”)) AND (LIMIT-TO (EXACTSRCTITLE, “Journal Of Nepal Health Research Council”)). The method resulted in the number of articles indexed under the journal title. For the purpose of this study, we excluded the ‘review articles’ and ‘editorials’ as editors may be obliged to write such articles to promote and pave the direction of the journals. In general editorial practice, editorials do not undergo a peer review process but the other article categories such as ‘full-length article’ (Scopus identifies it as “ar”) and ‘review article’ (Scopus identifies as “re”) should be peer-reviewed.

For the analysis of the data, we used social network analysis applications. VOSviewer was used to visualise the authors who contributed to the journals. Since the Scopus database identifies authors by surname (as the first name) followed by the initials of the first and middle names, it posed a problem of multiple authors being counted as a single author. To avoid this possibility, we used the Scopus author identification numbers – the unique numbers assigned to each author - believing that identifying authors by Scopus identification numbers is more accurate than by the authors’ last names and initials.

## Results

### Predominance of Select Authors

Over the years, down to 2021, some of the six journals analysed have increased the number of published articles exponentially. JNMA was established in 1963 but got indexed in Scopus in 2005 only when it published 28 articles. The number increased by 825 per cent to reach 259 articles in 2021. Along with the increase in the number of articles, the journal provided a significant percentage of publication space to selective authors only. For example, an author published seven articles (2.7 per cent of the total articles) in 2021, another seven (2.89 per cent) in 2020 and five articles (4.13 per cent) in 2019. In 2007, it had provided 8.57 per cent of publication space to an individual author.

**Table 2.** Predominance of Select authors in a selected Nepali Medical Journals

| Year | NHRC |  |  | KUMJ |  |  | NMCJ |  |  | JNPS |  |  | NJO |  |  | JNMA |  |  |
| --- | --- | --- | --- | --- | --- | --- | --- | --- | --- | --- | --- | --- | --- | --- | --- | --- | --- | --- |
|  | Total Articles | Maximum no of articles by single author | Percentage | Total Articles | Maximum no of articles by single author | Percentage | Total Articles | Maximum no of articles by single author | Percentage | Total Articles | Maximum no of articles by single author | Percentage | Total Articles | Maximum no of articles by single author | Percentage | Total Articles | Maximum no of articles by single author | Percentage |
| 2021 | 131 | 8 | 6.11% | 57 | 4 | 7.02% | - | - | - | 50 | 3 | 6.00% | - | - | - | 259 | 7 | 2.70% |
| 2020 | 143 | 15 | 10.49% | 116 | 9 | 7.76% | - | - | - | 44 | 3 | 6.82% | - | - | - | 242 | 7 | 2.89% |
| 2019 | 105 | 8 | 7.62% | 72 | 6 | 8.33% | - | - | - | 35 | 2 | 5.71% | - | - | - | 121 | 5 | 4.13% |
| 2018 | 68 | 7 | 10.29% | 79 | 4 | 5.06% | - | - | - | 57 | 3 | 5.26% | - | - | - | 103 | 4 | 3.88% |
| 2017 | 37 | 2 | 5.41% | 82 | 5 | 6.10% | - | - | - | 40 | 2 | 5.00% | - | - | - | 71 | 4 | 5.63% |
| 2016 | 41 | 3 | 7.32% | 92 | 5 | 5.43% | - | - | - | 70 | 2 | 2.86% | 27 | 2 | 7.41% | 42 | 2 | 4.76% |
| 2015 | 45 | 9 | 20.00% | 58 | 5 | 8.62% | - | - | - | 62 | 3 | 4.84% | 16 | 4 | 25.00% | 77 | 3 | 3.90% |
| 2014 | 45 | 2 | 4.44% | 73 | 4 | 5.48% | 48 | 4 | 8.33% | 54 | 3 | 5.56% | 17 | 4 | 23.53% | 63 | 2 | 3.17% |
| 2013 | 63 | 6 | 9.52% | 72 | 10 | 13.89% | 38 | 3 | 7.89% | 61 | 3 | 4.92% | 51 | 2 | 3.92% | 83 | 3 | 3.61% |
| 2012 | 33 | 2 | 6.06% | 79 | 6 | 7.59% | 82 | 8 | 9.76% | 57 | 2 | 3.51% | 55 | 3 | 5.45% | 40 | 3 | 7.50% |
| 2011 | 41 | 4 | 9.76% | 65 | 4 | 6.15% | 80 | 9 | 11.25% | 38 | 3 | 7.89% | 37 | 4 | 10.81% | 33 | 2 | 6.06% |
| 2010 | 28 | 2 | 7.14% | 71 | 4 | 5.63% | 66 | 3 | 4.55% | 21 | 2 | 9.52% | 24 | 5 | 20.83% | 71 | 3 | 4.23% |
| 2009 | - | - | - | 75 | 3 | 4.00% | 75 | 5 | 6.67% | 18 | 4 | 22.22% | 22 | 4 | 18.18% | 72 | 3 | 4.17% |
| 2008 | - | - | - | 84 | 3 | 3.57% | 62 | 6 | 9.68% | 16 | 3 | 18.75% | - | - | - | 54 | 3 | 5.56% |
| 2007 | - | - | - | 104 | 5 | 4.81% | 67 | 6 | 8.96% | - | - | - | - | - | - | 35 | 3 | 8.57% |
| 2006 | - | - | - | 94 | 5 | 5.32% | 75 | 4 | 5.33% | - | - | - | - | - | - | 41 | 3 | 7.32% |
| 2005 | - | - | - | 86 | 5 | 5.81% | 41 | 3 | 7.32% | - | - | - | - | - | - | 28 | 2 | 7.14% |
| 2004 | - | - | - | 62 | 5 | 8.06% | 39 | 5 | 12.82% | - | - | - | - | - | - | - | - | - |
| 2003 | - | - | - | 55 | 10 | 18.18% | 33 | 4 | 12.12% | - | - | - | - | - | - | - | - | - |

JNPS published 16 articles in 2008 – the number increased by over 212 per cent to 50 in the year 2021 – allowed a single author to publish four articles (occupying 22.22 per cent of the annual publication space) in 2009. Likewise, KUMJ published 55 articles in 2003, which increased to 57 in the year 2021. It provided 13.89 per cent of the journal space for a single author to publish in 2013, allowing an author to publish 10 out of 72 published articles. In 2010, NHRC had 28 articles, which reached 131 in 2021, with an increase of 368 per cent. In 2015, a single author published nine (20 per cent) out of 45 articles in the NHRC journal and 15 (10.49 per cent) articles out of 143 published articles by a single author in 2020. NJO, which was removed from Scopus indexing in 2016, published 22 articles in 2009. Seven years down the line, in 2016, the number decreased to 16. In 2015, it also allowed a single author to publish four articles (25 per cent) of the total articles. NMCJ, another journal removed from the Scopus registration, published 33 articles in 2003. It was indexed in Scopus until 2014 when it published 48 articles. In 2004, it provided 12.82 per cent of the space for a single author allowing them to publish five articles.

### Co-authorship Networks

Co-authorship information revealed that those working as nodal co-authors were also found to be the editors of journals. Across all journals of our study, 109 authors were found to have published articles in the journals as editors. Twenty-two editorial board members have published at least one article between 2017 and 2021 and the highest number of articles by a single editorial board member of the NHRC journal during that period is 40. Similarly, six editorial board members of KUMJ have published at least one article between 2017 and 2021, with the highest number of single-author articles being 22. In the JNMA journal, 21 editorial board members have published at least one article during 2017 – 2021, with an individual authoring as high as 11 articles. Eight editorial board members of NMCJ journal had published at least one article during 2010 to 2014 (last five years before it was delisted), with one member published 24 articles. The JNPS journal publishes a small number of articles compared to other journals annually, however, it also follows similar trends of editorial involvement. In our dataset, we found that one editor published four articles from 2017 to 2021. The NJO is not an exception. Fourteen editorial members had published at least one article from 2012 to 2016 (five years before it was delisted) and one of them published six articles during that period.

**Figure 1.**
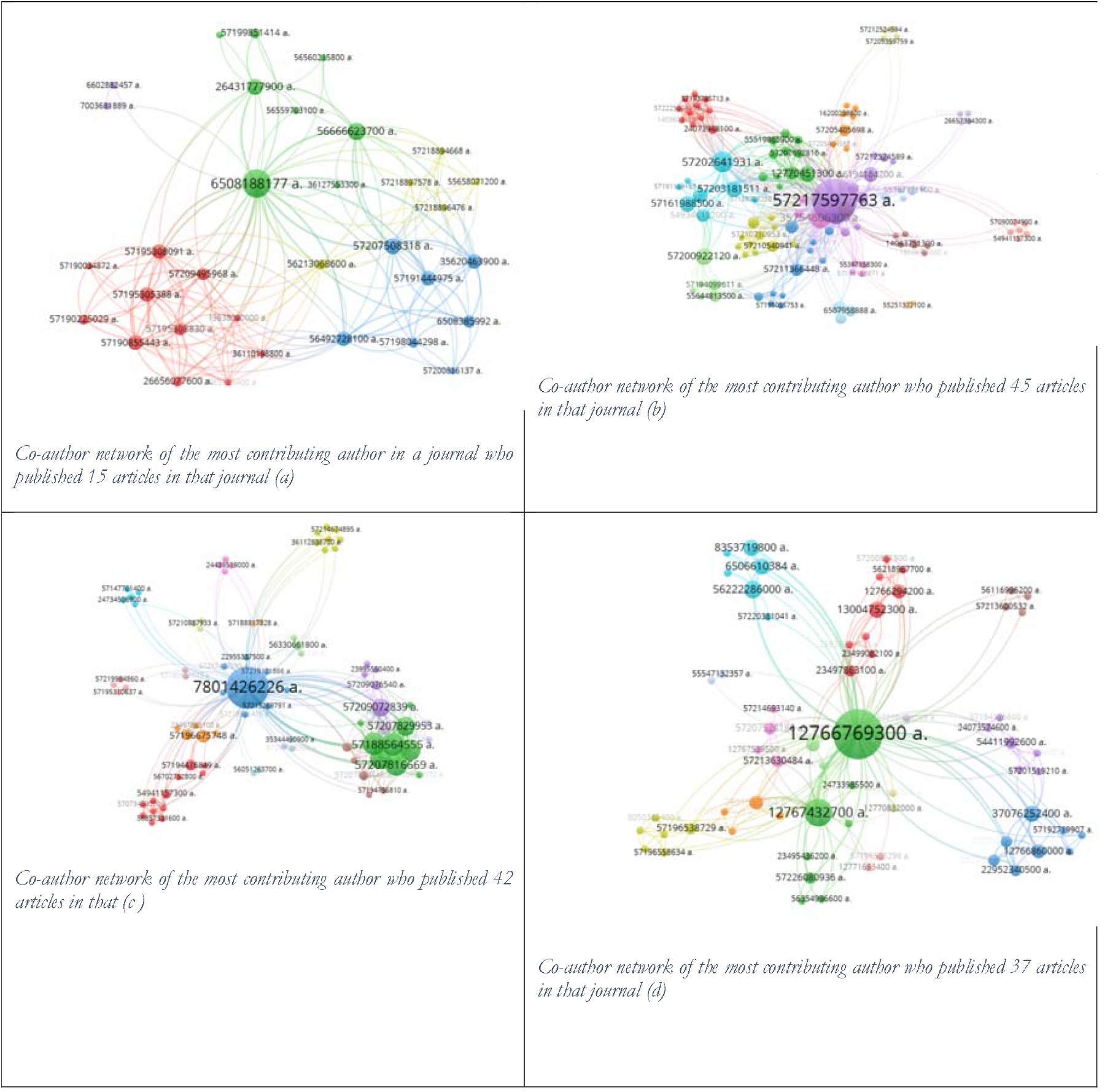
Co-authorship network of major node (a, b, c are editor of the journal, d is not)

**Table 1.** Maximum Percentage of articles where the individual editor appeared as a co-author each year

| Year | JNMA | NHRC | KUMJ | NMCJ | JNPS | NJO |
| --- | --- | --- | --- | --- | --- | --- |
| 2021 | 1.54% | 6.11% | 7.02% | - | 4.00% | - |
| 2020 | 2.48% | 10.49% | 6.90% | - | 2.27% | - |
| 2019 | 1.65% | 7.62% | 8.33% | - | 2.86% | - |
| 2018 | 3.88% | 10.29% | 3.80% | - | 0 | - |
| 2017 | 2.82% | 5.41% | 2.44% | - | 0 | - |
| 2016 | 2.38% | 7.32% | 5.43% | - | 0 | 7.41% |
| 2015 | 3.90% | 20.00% | 8.62% | - | 0 | 6.25% |
| 2014 | 3.17% | 4.44% | 5.48% | 8.33% | 0 | 11.76% |
| 2013 | 2.41% | 4.76% | 13.89% | 5.26% | 1.64% | 3.92% |
| 2012 | 2.50% | 3.03% | 7.59% | 9.76% | 0 | 3.64% |
| 2011 | 0 | 9.76% | 4.62% | 11.25% | 2.63% | 5.41% |
| 2010 | 2.82% | 3.57% | 4.23% | 3.03% | 4.76% | 12.50% |
| 2009 | 2.78% | - | 1.33% | 6.67% | 0 | 18.18% |
| 2008 | 3.70% | - | 1.19% | 9.68% | 6.25% | - |
| 2007 | 2.86% | - | 0.96% | 2.99% | - | - |
| 2006 | 4.88% | - | 3.19% | 5.33% | - | - |
| 2005 | 0 | - | 2.33% | 4.88% | - | - |
| 2004 | - | - | 8.06% | 12.82% | - | - |
| 2003 | - | - | 5.45% | 12.12% | - | - |

## Discussion and Conclusion

Journals in a small scientific community serve an important educational function, and they provide outlets for researchers to learn research reporting and report specific local knowledge. Because of the function, a smaller number of authors are expected to publish more articles in journals. This may not be true for the editorial board members as they are the quality controller of the journals and also serve as the knowledge bridge between mainstream academic journals and national journals. However, it is important to differentiate between the publications in a journal by editors having a decision-making role, and the others with other functions such as reviewing and writing articles. The conflict of interest of the decision-making editors differs from that of the other researchers involved in a journal. This is why there are standards and best practices to deal with the conflict of interest of decision-making editors, such as WAME policies. It encourages research on the principles and practice of medical editing (see www.wame.org), among others.

The dominance of a few authors is a feature that cuts across all the medical journals we have reviewed. An editor who published 57 articles was found to publish as many as 45 articles in one of the journals he edited. A large number of articles by a single author in the journals over the years may not be a problem, provided that the author also contributes with their articles in other international journals. Multiple research publications by a scholar in any journal indicate their productivity and, as such, should not be an issue (Abbasi et al., 2011). The fit, quality, and speed of publication determine authors’ choice of a journal (Solomon & Björk, 2012). We found that some contributors have exclusively contributed to the journals whereas there exist many other with similar aims and scopes. It provides a ground to doubt that they may have been preferentially treated. The authors do not contribute to other local journals either. Hence, allowing unusual publishing space for a single author in Nepali medical journals is worrying. It reflects the biased closeness of the community in question and promotes scientific inbreeding whereby a certain groups of professionals have exclusively promoted themselves, barring the other contributors from using the outlets to share their knowledge products, or for the other contributors the close-community is an alien space. This reality raises the question of editorial transparency and provides enough space to suspect why a particular author decides a specific journal to publish multiple research works in the same year.

We found that two of the journals we studied were delisted from the Scopus database. Presumably, failure to comply with the Scopus standard could be the reason for getting delisted. If the current trend of preferential and sequestered publication continues, other journals too run the risk of being delisted from Scopus. Also, unless we pay due attention to minimizing preferential publication biases, it will be difficult for the other journals to get listed in the international quality indexing system. If that happens, the chances of these non-mainstream journals contributing to hybrid knowledge as trusted partners of mainstream journals will be over. The delisting also questions the validity, quality and standard of the knowledge generated and disseminated to date thereby posing a serious question – what will be the fate and future of the other medical journals of Nepal that showcase Nepal’s medical education? Keeping on enjoying the status-quo and failing to comply with the established editorial practice will be a worrisome situation for the sector.

The six journals are the front face of Nepal’s medical education and research. The quality and rigour they maintain uphold their professional integrity and elevate them as a trusted source of scientific knowledge, which mainstream journals can use without a doubt. They can, thus, be the medium to bridge the gap between local and global knowledge, and also an indicator that Nepal’s medical education is on par with the rest of the world. All it requires is for the editors and managers of the journals to take a bold step to overhaul the editorial and management practices.

Technical journals, such as those we have reviewed, also contribute – or ‘should’ contribute – to policies that relate to the everyday lives of the people at large. To rise to these expectations and disciplinary promises, they should not operate in silos and be under the control of a few individuals whatever outstanding track record they maintain. Ideas – and pieces of knowledge for that matter – are created and co-created collectively in participation of as many people as possible, both experts and novices, to address outstanding gaps between research and practice, which are as glaring in the medical field as in others (Langley et al., 2018), more so in Nepal, given a limited focus and investment in research. By way of conclusion, we suggest the following four areas for further investigation in order to arrive at the clarity and understanding of some of the issues we have flagged in our findings and discussions.

a. Self-promotion by editors via editor-authorship is one of the recurring features in the journals investigated. However, why it has happened was not the question we investigated, we recommend undertaking a focused study to explore the factors that underlie excess self-publication and its consequences, both short-term and long-term.
b. The reliance of authors on a particular journal is another. The authors have multiple outlets to pick and choose from. Why they, however, do rely on one particular outlet is an outstanding issue for further research and exploration. We leave it, too, for other researchers to examine.
c. What motivates the exclusive networking of authors – who actually benefit when going broad, inclusive and participatory – is yet another question that we think should be critically examined.

## Data Availability

All article information used in this study is available in Scopus database. Editorial information were collected from their respective journal websites. The procedure to collect article information from the Scopus has been clearly mentioned in the manuscript

http://www.scopus.com

http://www.nepjol.info

## Appendix A

Most Articles by a single author in Nepali Medical Journal (Total number of articles in Scopus by author is captured by search code (AU-ID (AuthorScopusID) AND PUBYEAR < 2022)

| Total Articles in Scopus by author | Total article in Nepalese Journal indexed in Scopus | JNMA | KUMJ | NHRC | NMCJ | JNPS | NJO | Valus is 1 if the author is Editor to the journal |  |  |  |  |  |
| --- | --- | --- | --- | --- | --- | --- | --- | --- | --- | --- | --- | --- | --- |
|  |  |  |  |  |  |  |  | JNMA | KUMJ | NHRC | NMCJ | JNPS | NJO |
| 85 | 59 | 6 | 11 | 42 | 0 | 0 | 0 | 1 | 0 | 1 | 0 | 0 | 0 |
| 57 | 48 | 1 | 45 | 2 | 0 | 0 | 0 | 0 | 1 | 0 | 0 | 0 | 0 |
| 54 | 44 | 5 | 37 | 0 | 1 | 0 | 1 | 0 | 0 | 0 | 0 | 0 | 0 |
| 56 | 33 | 11 | 12 | 1 | 8 | 0 | 1 | 0 | 0 | 0 | 0 | 0 | 0 |
| 32 | 30 | 7 | 18 | 3 | 2 | 0 | 0 | 0 | 0 | 0 | 0 | 0 | 0 |
| 32 | 29 | 11 | 12 | 6 | 0 | 0 | 0 | 0 | 0 | 0 | 0 | 0 | 0 |
| 40 | 28 | 1 | 25 | 2 | 0 | 0 | 0 | 0 | 0 | 0 | 0 | 0 | 0 |
| 34 | 27 | 1 | 23 | 1 | 2 | 0 | 0 | 0 | 0 | 0 | 1 | 0 | 0 |
| 27 | 25 | 6 | 16 | 3 | 0 | 0 | 0 | 0 | 0 | 0 | 0 | 0 | 0 |
| 33 | 25 | 5 | 18 | 2 | 0 | 0 | 0 | 0 | 0 | 0 | 0 | 0 | 0 |
| 28 | 25 | 0 | 23 | 1 | 1 | 0 | 0 | 0 | 1 | 0 | 0 | 0 | 0 |
| 30 | 24 | 3 | 20 | 0 | 1 | 0 | 0 | 0 | 0 | 0 | 0 | 0 | 0 |
| 36 | 24 | 2 | 5 | 2 | 1 | 14 | 0 | 0 | 0 | 0 | 0 | 0 | 0 |
| 23 | 23 | 12 | 8 | 3 | 0 | 0 | 0 | 1 | 0 | 0 | 0 | 0 | 0 |
| 29 | 23 | 3 | 3 | 8 | 0 | 9 | 0 | 0 | 0 | 0 | 0 | 0 | 0 |
| 36 | 22 | 2 | 20 | 0 | 0 | 0 | 0 | 0 | 0 | 0 | 0 | 0 | 0 |
| 23 | 22 | 1 | 19 | 0 | 1 | 1 | 0 | 0 | 0 | 0 | 0 | 0 | 0 |
| 25 | 21 | 1 | 20 | 0 | 0 | 0 | 0 | 0 | 0 | 0 | 0 | 0 | 0 |
| 36 | 21 | 2 | 19 | 0 | 0 | 0 | 0 | 0 | 0 | 0 | 0 | 0 | 0 |
| 40 | 21 | 2 | 16 | 2 | 1 | 0 | 0 | 0 | 0 | 0 | 0 | 0 | 0 |
| 26 | 21 | 1 | 17 | 0 | 3 | 0 | 0 | 0 | 0 | 0 | 0 | 0 | 0 |
| 39 | 21 | 0 | 0 | 0 | 21 | 0 | 0 | 0 | 0 | 0 | 0 | 0 | 0 |
| 22 | 21 | 6 | 9 | 5 | 0 | 1 | 0 | 0 | 0 | 0 | 0 | 0 | 0 |
| 26 | 21 | 3 | 8 | 1 | 0 | 9 | 0 | 0 | 0 | 0 | 0 | 0 | 0 |
| 22 | 20 | 6 | 8 | 6 | 0 | 0 | 0 | 0 | 0 | 0 | 0 | 0 | 0 |
| 24 | 20 | 1 | 18 | 0 | 1 | 0 | 0 | 0 | 0 | 0 | 0 | 0 | 0 |
| 20 | 20 | 0 | 18 | 0 | 2 | 0 | 0 | 0 | 0 | 0 | 0 | 0 | 0 |
| 39 | 20 | 0 | 12 | 1 | 7 | 0 | 0 | 0 | 0 | 0 | 0 | 0 | 0 |
| 31 | 20 | 4 | 3 | 1 | 11 | 0 | 1 | 0 | 0 | 0 | 1 | 0 | 0 |
| 22 | 18 | 11 | 6 | 1 | 0 | 0 | 0 | 0 | 0 | 0 | 0 | 0 | 0 |
| 19 | 18 | 2 | 16 | 0 | 0 | 0 | 0 | 0 | 0 | 0 | 0 | 0 | 0 |
| 23 | 18 | 2 | 16 | 0 | 0 | 0 | 0 | 0 | 0 | 0 | 0 | 0 | 0 |
| 23 | 18 | 1 | 13 | 2 | 2 | 0 | 0 | 0 | 0 | 0 | 0 | 0 | 0 |
| 19 | 18 | 7 | 7 | 2 | 2 | 0 | 0 | 0 | 0 | 0 | 0 | 0 | 0 |
| 32 | 18 | 12 | 4 | 0 | 2 | 0 | 0 | 0 | 0 | 0 | 0 | 0 | 0 |
| 22 | 18 | 0 | 10 | 0 | 0 | 8 | 0 | 0 | 0 | 0 | 0 | 0 | 0 |
| 19 | 17 | 2 | 3 | 12 | 0 | 0 | 0 | 0 | 0 | 0 | 0 | 0 | 0 |
| 22 | 17 | 1 | 12 | 4 | 0 | 0 | 0 | 0 | 1 | 0 | 0 | 0 | 0 |
| 19 | 17 | 0 | 15 | 2 | 0 | 0 | 0 | 0 | 0 | 0 | 0 | 0 | 0 |
| 20 | 17 | 2 | 14 | 1 | 0 | 0 | 0 | 0 | 0 | 0 | 0 | 0 | 0 |
| 27 | 17 | 11 | 3 | 1 | 2 | 0 | 0 | 0 | 0 | 0 | 0 | 0 | 0 |
| 32 | 17 | 1 | 6 | 6 | 4 | 0 | 0 | 0 | 0 | 0 | 0 | 0 | 0 |
| 27 | 17 | 2 | 5 | 1 | 9 | 0 | 0 | 0 | 0 | 0 | 0 | 0 | 0 |
| 33 | 17 | 4 | 0 | 2 | 3 | 7 | 1 | 0 | 0 | 0 | 0 | 0 | 0 |
| 24 | 16 | 4 | 0 | 12 | 0 | 0 | 0 | 1 | 0 | 1 | 0 | 0 | 0 |
| 17 | 16 | 1 | 12 | 3 | 0 | 0 | 0 | 0 | 0 | 0 | 0 | 0 | 0 |
| 24 | 16 | 1 | 14 | 1 | 0 | 0 | 0 | 0 | 0 | 0 | 0 | 0 | 0 |
| 18 | 16 | 0 | 16 | 0 | 0 | 0 | 0 | 0 | 0 | 0 | 0 | 0 | 0 |
| 22 | 16 | 1 | 15 | 0 | 0 | 0 | 0 | 0 | 0 | 0 | 0 | 0 | 0 |
| 28 | 16 | 15 | 1 | 0 | 0 | 0 | 0 | 0 | 0 | 0 | 0 | 0 | 0 |
| 26 | 16 | 5 | 8 | 1 | 2 | 0 | 0 | 0 | 0 | 0 | 0 | 0 | 0 |
| 18 | 16 | 3 | 10 | 2 | 0 | 1 | 0 | 0 | 0 | 0 | 0 | 0 | 0 |
| 24 | 16 | 7 | 3 | 4 | 1 | 1 | 0 | 0 | 0 | 0 | 0 | 0 | 0 |
| 26 | 15 | 7 | 1 | 7 | 0 | 0 | 0 | 1 | 0 | 0 | 0 | 0 | 0 |
| 15 | 15 | 11 | 0 | 4 | 0 | 0 | 0 | 1 | 0 | 0 | 0 | 0 | 0 |
| 17 | 15 | 9 | 2 | 4 | 0 | 0 | 0 | 0 | 0 | 0 | 0 | 0 | 0 |
| 15 | 15 | 9 | 4 | 2 | 0 | 0 | 0 | 0 | 0 | 0 | 0 | 0 | 0 |
| 18 | 15 | 0 | 11 | 3 | 1 | 0 | 0 | 0 | 0 | 0 | 0 | 0 | 0 |
| 19 | 15 | 6 | 5 | 3 | 1 | 0 | 0 | 0 | 0 | 0 | 0 | 0 | 0 |
| 18 | 15 | 10 | 3 | 1 | 1 | 0 | 0 | 0 | 0 | 0 | 0 | 0 | 0 |
| 24 | 15 | 6 | 6 | 1 | 2 | 0 | 0 | 0 | 0 | 0 | 0 | 0 | 0 |
| 21 | 15 | 7 | 6 | 0 | 2 | 0 | 0 | 0 | 0 | 0 | 0 | 0 | 0 |
| 16 | 15 | 6 | 4 | 1 | 4 | 0 | 0 | 0 | 0 | 0 | 0 | 0 | 0 |
| 21 | 15 | 3 | 1 | 1 | 10 | 0 | 0 | 0 | 0 | 0 | 0 | 0 | 0 |
| 16 | 15 | 1 | 11 | 0 | 0 | 3 | 0 | 0 | 0 | 0 | 0 | 0 | 0 |
| 20 | 15 | 0 | 11 | 0 | 1 | 3 | 0 | 0 | 0 | 0 | 0 | 0 | 0 |
| 23 | 15 | 3 | 4 | 0 | 0 | 0 | 8 | 0 | 0 | 0 | 0 | 0 | 0 |
| 15 | 14 | 0 | 2 | 12 | 0 | 0 | 0 | 0 | 0 | 0 | 0 | 0 | 0 |
| 15 | 14 | 0 | 2 | 12 | 0 | 0 | 0 | 0 | 0 | 0 | 0 | 0 | 0 |
| 14 | 14 | 0 | 3 | 11 | 0 | 0 | 0 | 0 | 0 | 0 | 0 | 0 | 0 |
| 23 | 14 | 2 | 5 | 7 | 0 | 0 | 0 | 1 | 0 | 0 | 0 | 0 | 0 |
| 17 | 14 | 10 | 2 | 2 | 0 | 0 | 0 | 0 | 0 | 0 | 0 | 0 | 0 |
| 19 | 14 | 5 | 9 | 0 | 0 | 0 | 0 | 0 | 0 | 0 | 0 | 0 | 0 |
| 15 | 14 | 5 | 9 | 0 | 0 | 0 | 0 | 0 | 0 | 0 | 0 | 0 | 0 |
| 14 | 14 | 7 | 2 | 2 | 3 | 0 | 0 | 0 | 0 | 0 | 0 | 0 | 0 |
| 14 | 14 | 2 | 7 | 1 | 4 | 0 | 0 | 0 | 0 | 0 | 0 | 0 | 0 |
| 14 | 14 | 3 | 1 | 0 | 10 | 0 | 0 | 0 | 0 | 0 | 1 | 0 | 0 |
| 14 | 13 | 1 | 12 | 0 | 0 | 0 | 0 | 0 | 1 | 0 | 0 | 0 | 0 |
| 14 | 13 | 0 | 1 | 0 | 12 | 0 | 0 | 0 | 0 | 0 | 1 | 0 | 0 |
| 16 | 11 | 1 | 10 | 0 | 0 | 0 | 0 | 0 | 1 | 0 | 0 | 0 | 0 |
| 11 | 10 | 9 | 1 | 0 | 0 | 0 | 0 | 1 | 0 | 0 | 0 | 0 | 0 |
| 13 | 10 | 1 | 4 | 0 | 5 | 0 | 0 | 0 | 1 | 0 | 0 | 0 | 0 |
| 10 | 10 | 0 | 5 | 4 | 0 | 0 | 1 | 0 | 0 | 0 | 0 | 0 | 1 |
| 10 | 10 | 1 | 1 | 0 | 7 | 0 | 1 | 0 | 0 | 0 | 1 | 0 | 0 |
| 13 | 10 | 0 | 2 | 0 | 1 | 1 | 6 | 0 | 0 | 0 | 0 | 0 | 1 |
| 19 | 10 | 1 | 0 | 1 | 0 | 0 | 8 | 0 | 0 | 0 | 0 | 0 | 1 |
| 16 | 9 | 1 | 7 | 1 | 0 | 0 | 0 | 0 | 1 | 0 | 0 | 0 | 0 |
| 12 | 8 | 4 | 0 | 4 | 0 | 0 | 0 | 0 | 0 | 1 | 0 | 0 | 0 |
| 8 | 8 | 5 | 0 | 3 | 0 | 0 | 0 | 1 | 0 | 0 | 0 | 0 | 0 |
| 11 | 8 | 0 | 8 | 0 | 0 | 0 | 0 | 0 | 1 | 0 | 0 | 0 | 0 |
| 9 | 8 | 2 | 6 | 0 | 0 | 0 | 0 | 1 | 0 | 0 | 0 | 0 | 0 |
| 11 | 7 | 1 | 0 | 6 | 0 | 0 | 0 | 0 | 0 | 1 | 0 | 0 | 0 |
| 8 | 7 | 6 | 0 | 1 | 0 | 0 | 0 | 1 | 0 | 0 | 0 | 0 | 0 |
| 11 | 7 | 2 | 5 | 0 | 0 | 0 | 0 | 0 | 0 | 0 | 0 | 0 | 1 |
| 7 | 7 | 7 | 0 | 0 | 0 | 0 | 0 | 1 | 0 | 0 | 0 | 0 | 0 |
| 10 | 6 | 5 | 0 | 1 | 0 | 0 | 0 | 1 | 0 | 0 | 0 | 0 | 0 |
| 6 | 6 | 6 | 0 | 0 | 0 | 0 | 0 | 1 | 0 | 0 | 0 | 0 | 0 |
| 7 | 6 | 6 | 0 | 0 | 0 | 0 | 0 | 1 | 0 | 0 | 0 | 0 | 0 |
| 6 | 6 | 6 | 0 | 0 | 0 | 0 | 0 | 1 | 0 | 0 | 0 | 0 | 0 |
| 7 | 6 | 3 | 0 | 2 | 0 | 1 | 0 | 1 | 0 | 0 | 0 | 0 | 0 |

## Appendix B

Co-author network formed from the article published in the particular journal. The VoS Viewer software was used to draw the co-author network where the edge represents the article published in a journal by at least two authors.

**Figure 2.**
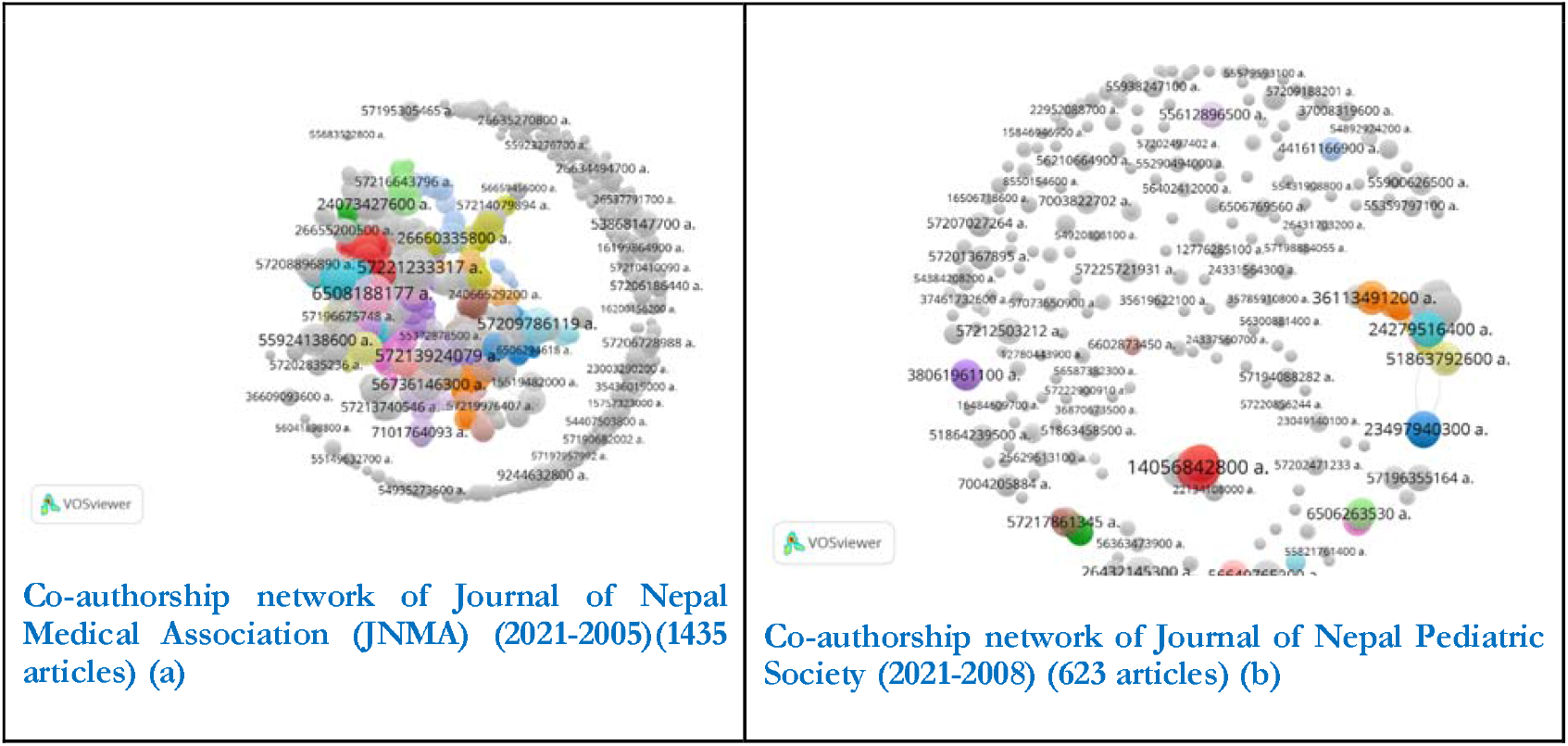
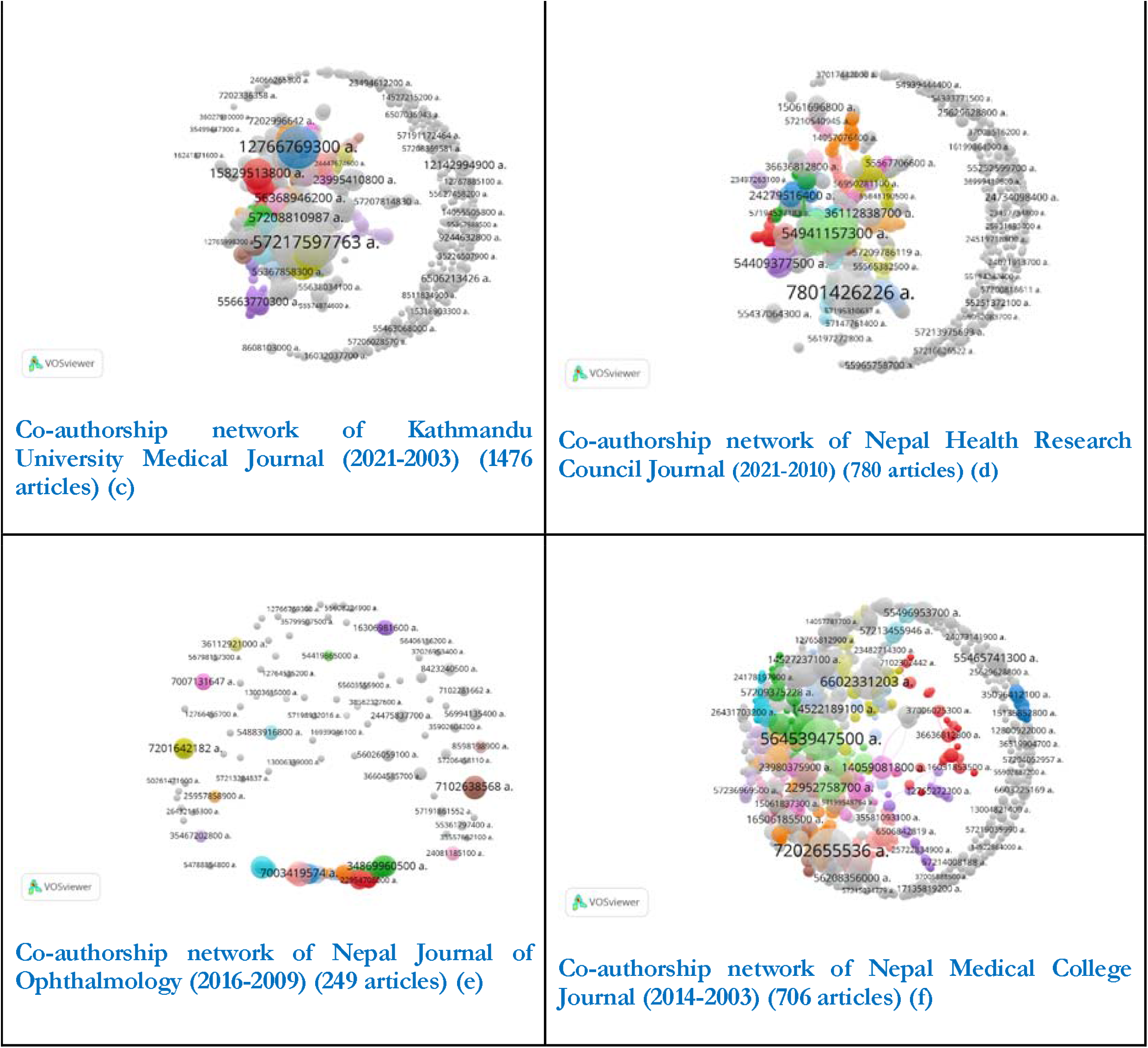
Network formed by the co-authors who published their articles in the corresponding Journal

